# Vision Language Model for Coronary Angiogram Analysis and Report Generation: Development and Evaluation Study

**DOI:** 10.64898/2026.04.19.26351241

**Authors:** Qianfeng Jiang, Yuhe Ke, Laura Gutierrez Sinisterra, Kabilan Elangovan, Zengxiang Li, Khung Keong Yeo, Yap Jonathan, Daniel Shu Wei Ting

## Abstract

Coronary artery disease is a leading cause of morbidity and mortality. Invasive coronary angiography is currently the gold standard in disease diagnosis. Several studies have attempted to use artificial intelligence (AI) to automate their interpretations with varying levels of success. However, most existing studies cannot generate detailed angiographic reports beyond simple classification or segmentation. This study aims to fine-tune and evaluate the performance of a Vision-Language Model (VLM) in coronary angiogram interpretation and report generation. Using twenty-thousand angiogram keyframes of 1987 patients collated across four unique datasets, we finetuned InternVL2-4B model with Low-Rank Adaptor weights that can perform stenosis detection, anatomy labelling, and report generation. The fine-tuned VLM achieved a precision of 0.56, recall of 0.64, and F1-score of 0.60 for stenosis detection. In anatomy segmentation, it attained a weighted precision of 0.50, recall of 0.43, and F1-score of 0.46, with higher scores in major vessel segments. Report generation integrating multiple angiographic projection views yielded an accuracy of 0.42, negative predictive value of 0.58 and specificity of 0.52. This study demonstrates the potential of using VLM to streamline angiogram interpretation to rapidly provide actionable information to guide management, support care in resource-limited settings, and audit the appropriateness of coronary interventions.

**AUTHOR SUMMARY:** Coronary artery disease has heavy disease burden worldwide and coronary angiogram is the gold standard imaging for its diagnosis. Interpreting these complex images and producing clinical reports require significant expertise and time. In this study, we fine-tuned and investigated an open-source VLM, InternVL2-4B, to interpret and report coronary angiogram images in key tasks including stenosis detection, anatomy identification, as well as full report generation. We also referenced the fine-tuned InternVL2-4B against state-of-the-art segmentation model, YOLOv8x, which was evaluated on the same test sets. We examined how machine learning metrics like the intersection over union score may not fully capture the clinical accuracy of model predictions and discussed the limitations of relying solely on these metrics for evaluating clinical AI systems. Although the model has not yet achieved expert-level interpretation, our results demonstrate the potential and feasibility of automating the reporting of coronary angiograms. Such systems could potentially assist cardiologists by improving reporting efficiency, highlightning lesions that may require review, and enabling automated calculations of clinical scores such as the SYNTAX score.

## 1 INTRODUCTION

Coronary artery disease (CAD), characterised by atherosclerotic plaques in coronary vessels, is a prevalent and leading cause of mortality both locally and worldwide [1]. Globally, the disease burden exceeds 7 million deaths annually [2]. Current practice involves recommending coronary angiography for patients identified as high risk based on risk factors or symptoms [3]. Coronary angiograms play a significant role in diagnosing and assessing the severity of CAD and remain the gold standard for diagnosing the disease [4, 5]. The number of coronary angiograms performed annually exceeds a million in the United States [6]. In Singapore, the National Heart Centre performs about two thousand four hundred such cases in a year [7].

However, interpretation of angiograms can be challenging due to their complexity and variability, and often requires specialised training [5]. The volume of imaging performed, in addition to its complexities, places significant demands on clinicians [8]. In response, there has been a growing interest in deploying AI to improve clinical workflow, reduce the reading time of medical images, and improve diagnostic accuracy [9]. In recent years, several attempts have been made to utilise deep neural networks to automate the interpretation of coronary angiograms with some success. One of the prominent examples is CathAI, a deep-learning-based system built to analyse static coronary angiogram frames. Through a multiple-stage neural network pipeline, CathAI has attained considerable success in classifying angiographic projections, locating coronary arteries, and predicting the severity of stenoses [10]. DeepCoro, another deep-learning-based pipeline, took it further by utilising a video-based approach to process angiographic sequences instead of isolated frames and a Swin3D transformer that allows for quantitative grading of stenosis severity [11]. Despite their advancements, one major limitation of neural-network-based solutions lies in their outputs. While these models may be able to generate structured numerical reports, they lack natural language interpretability. Clinically, coronary angiogram reports also contain descriptive findings which contribute to a comprehensive assessment of the patient’s condition. For instance, in addition to documenting the presence of a lesion in a specific coronary artery segment, cardiologists may also describe whether the same lesion involves the ostium of an adjacent branch. Other important descriptors include anatomical anomalies and the presence of collaterals. Most systems based on neural networks or vision transformer architectures seldom capture such narrative nuances which can be clinically important. This gap can potentially be filled by VLMs, which are designed to integrate language understanding and generation abilities of Large Language Models with those of image analysis. A recent review of generative AI has also highlighted that foundation models including VLMs can be fine-tuned with domain-specific clinical data to achieve comparable performance to state-of-the-art traditional deep learning methods [12], thereby underscoring the potential of such an approach to interpreting coronary angiograms.

Previously, enormous progress has been made to leverage AI for medical report generation [13]. A review of VLMs in medical image analysis has shown that they are able to perform a wide range of tasks, including classification, segmentation, question-answering, and report generation [14]. Medical report generation using VLMs is best established in chest radiography, where several studies [15, 16] have trained VLMs to report chest X-rays based on open-source datasets such as IU X-RAY and MIMIC CXR. In Cardiology, comparable efforts are newer; a VLM named EchoCLIP has achieved remarkable performance in interpreting echocardiograms like detecting cardiac chamber dilations [17, 18]. The aim of this study is to utilise VLMs in coronary angiogram analysis, taking into account multiple projection views of coronary angiography [19] to generate a fully consolidated report. Leveraging VLMs to automate angiogram analysis could help cardiologists in real-time visual assessment during the procedures and streamline post-procedure report generation, reducing both time and clinical load. It could also improve patient safety by identifying stenoses potentially overlooked by clinicians. A VLM capable of accurately detecting, localising and grading stenosis severity could potentially be further developed to automate the computation of the Synergy Between Percutaneous Coronary Intervention With Taxus and Cardiac Surgery (SYNTAX) score, a widely used clinical tool in interventional cardiology which can be time-consuming to calculate [20].

This study aims to fine-tune and evaluate a VLM in performing key coronary angiogram tasks including stenosis detection and coronary artery anatomy segmentation. Additionally, we seek to investigate its ability to generate a full structured text report from multiple angiographic images acquired from different projection angles. We hypothesise that the current state of the art VLMs can achieve performance comparable or even exceeding that of conventional methods like neural networks.

## 2 MATERIALS AND METHODS

### 2.1 The Need for Fine-Tuning

While most VLMs are trained on extensive image-text data, several studies [21–23] have highlighted that most models often underperform in task-specific applications despite their impressive capabilities in general domain. This is in line with our preliminary testing on InternVL2-4B [24, 25], an open-source general-purpose VLM capable of multi-image analysis, which correctly identified an image as a coronary angiogram, but struggled to recognise left versus right coronary arteries and performed poorly in localising vessel stenosis. This finding underscored the need for domain-specific fine-tuning, especially in coronary angiography interpretation where complex anatomies and multiple projection angles are involved.

### 2.2 Overview of Fine-Tuning

The tasks in coronary angiogram interpretation and report generation can be categorised in increasing order of difficulty: keyframe selection, stenosis detection, anatomy segmentation, and text report generation. The keyframe selection task is achieved by fine-tuning a ViT, Google vit-large-patch32-384 [26], pretrained with ImageNet, to select keyframes from coronary angiogram videos (section 2.4). The remaining tasks (sections 2.5 - 2.7) are accomplished by finetuning InternVL2-4B on selected datasets (section 2.3).

### 2.3 Datasets

In this study, we utilised publicly available datasets and a proprietary dataset. The Coronary Artery Disease Invasive Coronary Angiography (CADICA) [27], Angiographic Dataset for Stenosis Detection (ADSD) [28] and Automatic Region-based Coronary Artery Disease diagnostics using X-ray angiography imagEs (ARCADE) [29] datasets are open-source. These datasets contain expert-selected angiogram keyframes and bounding box coordinates of stenosis or coronary artery anatomy at individual image levels. The Artificial Intelligence in Medicine Transformation (AIMx) dataset is a proprietary dataset prospectively collected by the National Heart Centre Singapore from June 2021 to June 2024 (Protocol No. R1808/50/2021, CIRB Ref No. 2021/2518), where patients who were referred for angiography were recruited. All enrolled patients provided informed consent, with all methods performed in accordance with the relevant guidelines and regulations. The dataset was analysed retrospectively in this study. The AIMx dataset includes raw angiogram videos, and the diagnostic text report of angiogram findings for each patient. An overview of the datasets is shown in Table 1.

**Table 1.** Overview of datasets.

| Dataset | CADICA | ADSD | ARCADE (Stenosis) | ARCADE (Anatomy) | AIMx |
| --- | --- | --- | --- | --- | --- |
| Institution & Country | Hospital Universitario Virgen de la Victoria, Málaga, Spain | Research Institute for Complex Problems of Cardiovascular Diseases, Kemerovo, Russia | Research Institute of Cardiology and Internal Diseases, Kazakhstan |  | National Heart Centre Singapore, Singapore |
| Source | Open-source |  |  |  | Proprietary |
| Type of Ground Truth | Expert-annotated bounding box coordinates of stenosis |  |  | Expert-annotated bounding box coordinates of coronary artery segments according to SYNTAX definition | Consolidated text report of coronary angiogram findings |
| No. of Patients | 42 | 100 | 1500 |  | 345 |
| No. of Selected Videos | 382 | Information Unavailable |  |  | 3118 |
| No. of Keyframes | 6126 | 8326 | 1500 | 1500 | N/A |
| No. of Non-Keyframe | 13,346 | 0 | 0 | 0 | N/A |
| No. of Instances of Stenosis | 6161 | 8326 | 2030 | N/A | N/A |
| Area of Stenosis Bounding Box (Mean pixel<sup>2</sup>± SD) | 1635 ± 1367 | 674 ± 548 | 3476 ± 4300 | N/A | N/A |
| No. of Frames with No Stenosis | 2130 | 0 | 0 | N/A | N/A |
| Keyframe Selection | Pre-selected |  |  |  | Requires selection |

### 2.4 Keyframe Selection

Each coronary angiographic video begins with pre-injection frames in which the vessels are not visible under X-ray. As the contrast dye is introduced, the vessels progressively opacify, delineating their structures for diagnostic evaluation. The sequences conclude after the dissipation of the dyes. As such, only a subset of the frames contains diagnostic information per angiographic video. To reduce computational load and optimise efficiency, keyframe selection is an important task to only extract frames containing diagnostic information. While there is no universally standardised definition for keyframes, several studies have established heuristic criteria for identifying these frames. These criteria include vessel clarity, complete contrast filling of the vessels with visibility of both proximal and distal segments, and vessels being confined fully within the frame [30] (Table S1). Initial attempts utilising traditional algorithm-based selection methods such as Frangi filters [31, 32] did not yield satisfactory results on the AIMx dataset. The approach was susceptible to noise caused by curved structures such as ribcages and catheters, leading to the erroneous selection of non-keyframes. As such, a machine learning approach of training a keyframe selection model was adopted to help select keyframes from the AIMx dataset. The open-source datasets already contain pre-selected keyframes. The CADICA dataset also provided non-keyframes that do not satisfy the criteria above, which this study has also utilised for training the ViT. To incorporate some images from the AIMx dataset into training, 5 patients’ angiogram frames under the AIMx dataset were manually classified into keyframes and non-keyframes by a senior medical student. The fine-tuned ViT model was tested on 2 other patients’ angiogram images, which were also manually labelled by the same student. The Google vit-large-patch32-384 pretrained model [26] was selected for this task. It was fine-tuned and evaluated according to the breakdown in Fig S1 for 5 epochs using an NVIDIA RTX 4090 Graphical Processing Unit (GPU). The training batch size was set as 64 and the metric for evaluation was set as “accuracy”. All other training hyperparameters were set to the default in the PyTorch library. During inference, the ViT takes in each frame from the video and classifies it as either a keyframe or non-keyframe. Additionally, the ViT also provides a confidence score, which represents the probability that the frame belongs to the predicted category. Non-keyframes are defined as positives in this study to minimise false negative selections. The fine-tuned ViT was then used to run inferences on the test set, and its precision (positive predictive value), recall (sensitivity), accuracy, F1 score (harmonic mean of precision and recall) and AUC were computed.

### 2.5 Stenosis Detection

All images from the CADICA, ADSD and ARCADE datasets were resized to 448 × 448 pixels and converted to RGB format to be compatible for VLM fine-tuning. To ensure proportional representation of the datasets, image augmentation was applied 5 times for every training image in the ARCADE dataset. The augmentation included random rotations, elastic transformations, grid distortions, brightness-contrast adjustments and Gaussian blur.

An additional image processing function [29] was applied to enhance the contrast on all images. For evaluation, only the reserved test subset of 300 images from the ARCADE dataset was used, while training was conducted using CADICA, ADSD and the remaining from the ARCADE dataset. This decision was made because only the ARCADE dataset provided an official train-test split, which allows for a standardised comparison with other studies, in contrast to CADICA and ADSD, which do not have predefined train-test splits. Another reason for this decision is due to the limited variation of images in the CADICA and ADSD dataset, as many images were similar-looking to one another because they belonged to the same video of the same patient. This is in contrast to the ARCADE dataset, where the 1500 images were relatively unique.

In object detection tasks, the Intersection-over-Union (IoU) score, defined as the area of intersection between a predicted bounding box with that of the ground truth divided by their area of union, is a standard metric used to measure a model’s performance [33]. In this study, an IoU of 0.5 was set as the threshold. Each ground truth bounding box was matched against the bounding box predicted by the VLM. If the IoU exceeds 0.5, it would be considered a true positive (TP), and the ground truth will be marked as matched. If a predicted box has 60% of its area overlapping with that of a ground truth box, it will also be considered a TP. This additional step is not a standard practice in the evaluation of segmentation performance in AI models. However, in stenosis detection, the general location of the bounding box is more critical than the degree of overlap between predicted and ground truth box overlap. Including this step in the evaluation could potentially avoid misclassifying prediction as a false positive when the predicted box is much smaller than the ground truth box but is correctly identifying the location of stenosis. Predicted boxes not satisfying the above criteria were recorded as false positives (FP). For ground truth boxes unmatched by any predicted box, they were considered false negatives (FN). Each ground truth can only be matched once to prevent overcounting positive predictions which will inflate the model’s performance. The precision, recall and the F1 score were then computed for the consolidated outputs across all test images.

### 2.6 Anatomy Labelling

The ARCADE (anatomy) dataset contains 1500 images with bounding box coordinates of coronary artery segments annotated according to the SYNTAX definition [29]. Image augmentation was performed 5 times on each of the 1200 training images. They were then resized and processed similarly to what was performed on the ARCADE (stenosis) dataset. The official test subset of 300 images was used for evaluation. As the location and area of prediction of each artery segment are clinically significant, only the IoU threshold of 0.5 was used as the evaluation criteria, without the 60% overlap consideration, in contrast to the stenosis detection task. Fig S2 shows the number of instances of each vessel segment in the training dataset before image augmentation.

The vessel abbreviations and vessel segment designations used in Fig S2 are mapped to their SYNTAX nomenclature in Table S2.

### 2.7 Report Generation

The AIMx dataset comprises angiogram videos and diagnostic text reports from 345 patients. 305 patients’ data were randomly assigned for training, while the remaining were reserved for testing. All angiogram videos were manually reviewed, and peri-intervention and post-intervention videos were excluded, retaining only diagnostic angiogram videos. Angiograms demonstrating previously stented vessels and chronic total occlusions were included, while those involving graft vessels were excluded, with corresponding textual descriptions of graft vessel patency removed from the reports.

The fine-tuned frame selection ViT (section 2.4) was employed to extract keyframes. The frame with the highest confidence score was selected as the representative keyframe for every video. Each training sample corresponds to data from a single patient, consisting of one selected keyframe from each angiographic video, collectively paired with their diagnostic findings in text report format.

The VLM’s report generation performance was evaluated manually by comparing the findings in the model output to the ground truth report. Stenosis severity was classified into three main categories:

- None to mild: 0 – 49%
- Moderate: 50 – 70%
- Severe: > 70%, including total chronic occlusions

Moderate to severe stenoses were considered clinically significant [34]. Every VLM-generated statement was compared to the corresponding statement in the ground truth using the following criteria.

- If the VLM output a clinically significant stenosis (moderate or severe) that falls into the same category as the ground truth for a given vessel segment, it is considered a TP. Otherwise, it is counted as FP if it overstates the severity and FN if it understates the severity.
- If the VLM correctly identified a vessel segment with no to mild stenosis, it is considered a TN. Otherwise, it was counted as a FN. Vessel segments not described in the ground truth are assumed to have no stenosis for comparison purposes.
- Clinically significant stenoses in the ground truth not mentioned by the VLM output would be regarded as FN. Clinically insignificant ones would be ignored.

The accuracy, precision, recall, specificity and negative predictive value were then computed based on the collective findings in the test set.

### 2.8 Model Selection & Training Pipeline

As the report generation requires interpretation of multiple images to generate a consolidated paragraph of findings in a single input, a model that supports multi-image input was required. InternVL [24, 25], which supports fine-tuning with multiple images in a single input, was appropriate for this task. The training data was formatted as described in sections 2.5 – 2.7 and was fine-tuned on InternVL2-4B in a single setting. The vision backbone, multi-layer perceptron, and the language components were unfrozen during training. The fine-tuning was done on Low-Rank Adaptor (LoRA) weights, which reduces the computational complexity and allows for training on 2 NVIDIA RTX 4090 GPUs. The full training pipeline is illustrated in Fig 1, and was trained for a total of 5 epochs. The training parameters were set as the default according to InternVL’s repository, with some changes specified in Table S3.

**Fig 1.**
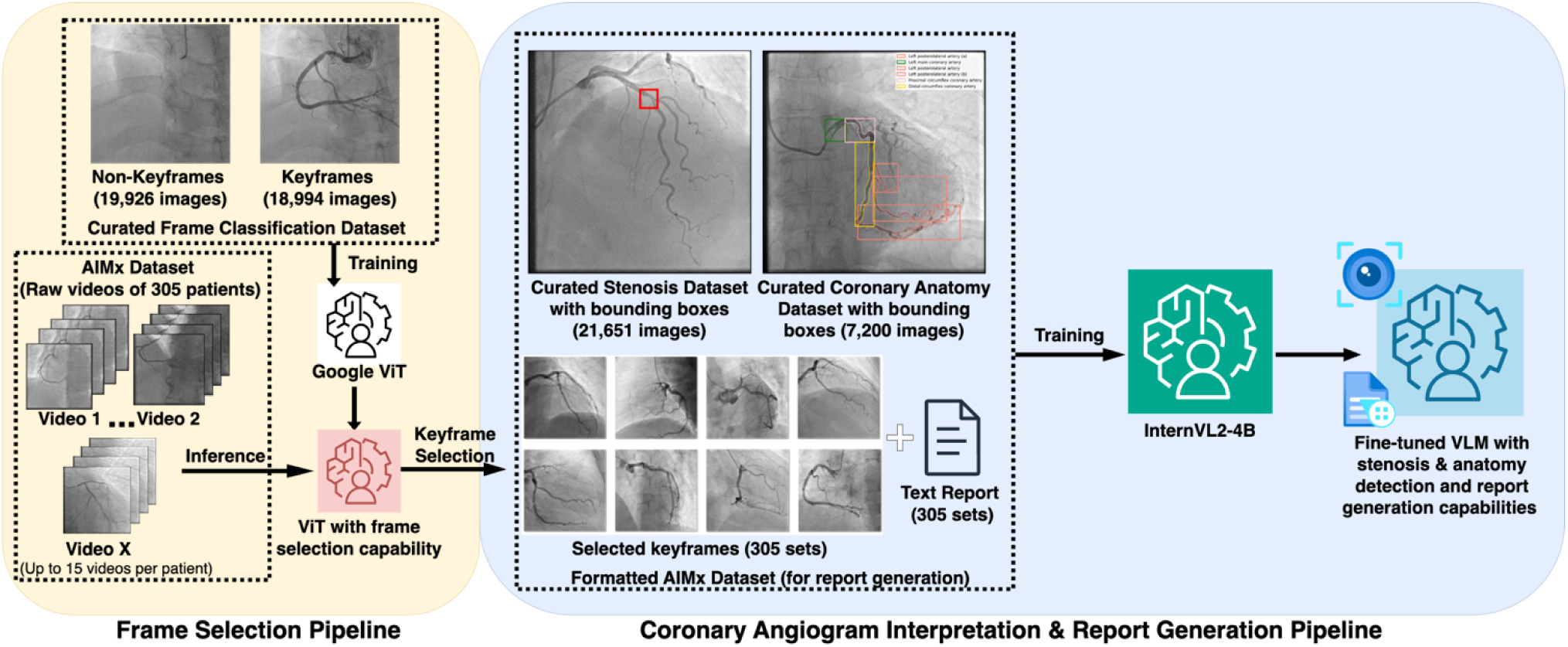
Overview of the full training pipeline for InternVL2-4B. A ViT was first fine-tuned to differentiate keyframes from non-keyframes, and subsequently used to infer and select one keyframe from each raw angiogram video in the AIMx dataset. The selected keyframes were then paired with the corresponding angiogram text report at the patient level and combined with the curated stenosis and anatomy datasets for fine-tuning of InternVL2-4B with LoRA.

## 3 RESULTS

### 3.1 Keyframe Selection

The fine-tuned ViT frame selector achieved TP = 1506, TN = 297, FP = 109 and FN = 179 on the 2091 test images. This gives an accuracy of 0.86, precision of 0.89, recall of 0.93, F1 score of 0.91 and AUC of 0.94 (Fig 2).

**Fig 2.**
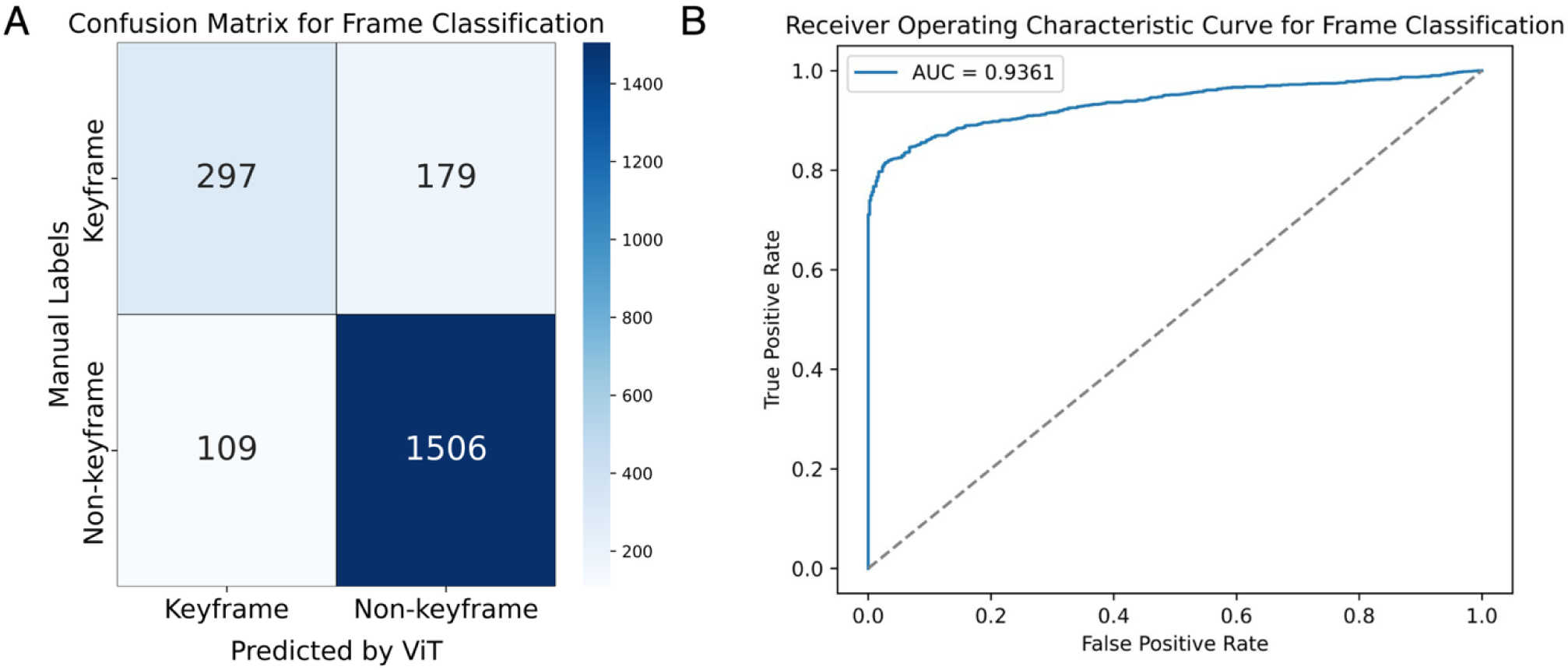
(A) Confusion matrix for keyframe classification. (B) Receiver operating characteristic curve for frame classification.

The ViT frame selector has achieved robust performance as it could recognise poor frames with high precision and recall. This is critical in the data pipeline for the subsequent VLM fine-tuning for report generation, as mistakenly selecting a non-keyframe with no diagnostic information could introduce noise into training data and affect downstream training. In our case, most errors arose from misclassifying a keyframe as a non-keyframe, suggesting that it is overly cautious on keyframe identification. However, this impact is minimal in our use case because we select only one frame with the highest confidence from > 100 frames per video. Additionally, the ViT has shown to be more reliable than algorithm-based approaches on the AIMx dataset, although a quantitative comparison was not made.

### 3.2 Stenosis Detection

The fine-tuned InternVL-4B VLM obtained a precision of 0.56, recall of 0.64 and F1-score of 0.60 on the test dataset (Table S4). As a reference, the performance achieved by YOLOv8x, a specialised segmentation model with a neural network architecture [35] which was fine-tuned with ARCADE training data, reported precision, recall and Dice scores of 0.36, 0.45 and 0.40 respectively on the same set of test images [29]. However, definitions of TP, FP and FN differ among our studies. YOLOv8x’s predictions were evaluated on a pixel level and ours were assessed on a bounding-box level. This makes our results not directly comparable. Nonetheless, this study has demonstrated that VLMs can perform stenosis detection tasks on par with neural network-based architectures.

### 3.3 Anatomy Labelling

Our VLM achieved a weighted average precision, recall and F1 score of 0.50, 0.43 and 0.46 respectively on the ARCADE anatomy test set. It performed better in the larger vessel segments, such as the left main, and proximal left circumflex arteries, with some scores exceeding 0.7 (Fig 3). However, it performed poorly in the smaller branches such as the obtuse marginal and diagonal arteries, some of which had scores of 0. This is an expected finding given that these larger vessel segments appeared much more frequently in the training dataset (Fig S2) and their anatomies were less complex. Vessel segments that were better represented, such as the main segments of the right coronary artery (RCA), also had higher scores. It was also observed that some distal vessel segments tend to have lower scores than their proximal counterparts. Our findings were in line with studies that used neural networks evaluated on the same test dataset, as they also reported better segmentation results with these larger and more common vessel segments, and extremely low scores on the less represented ones [29, 36, 37].

**Fig 3.**
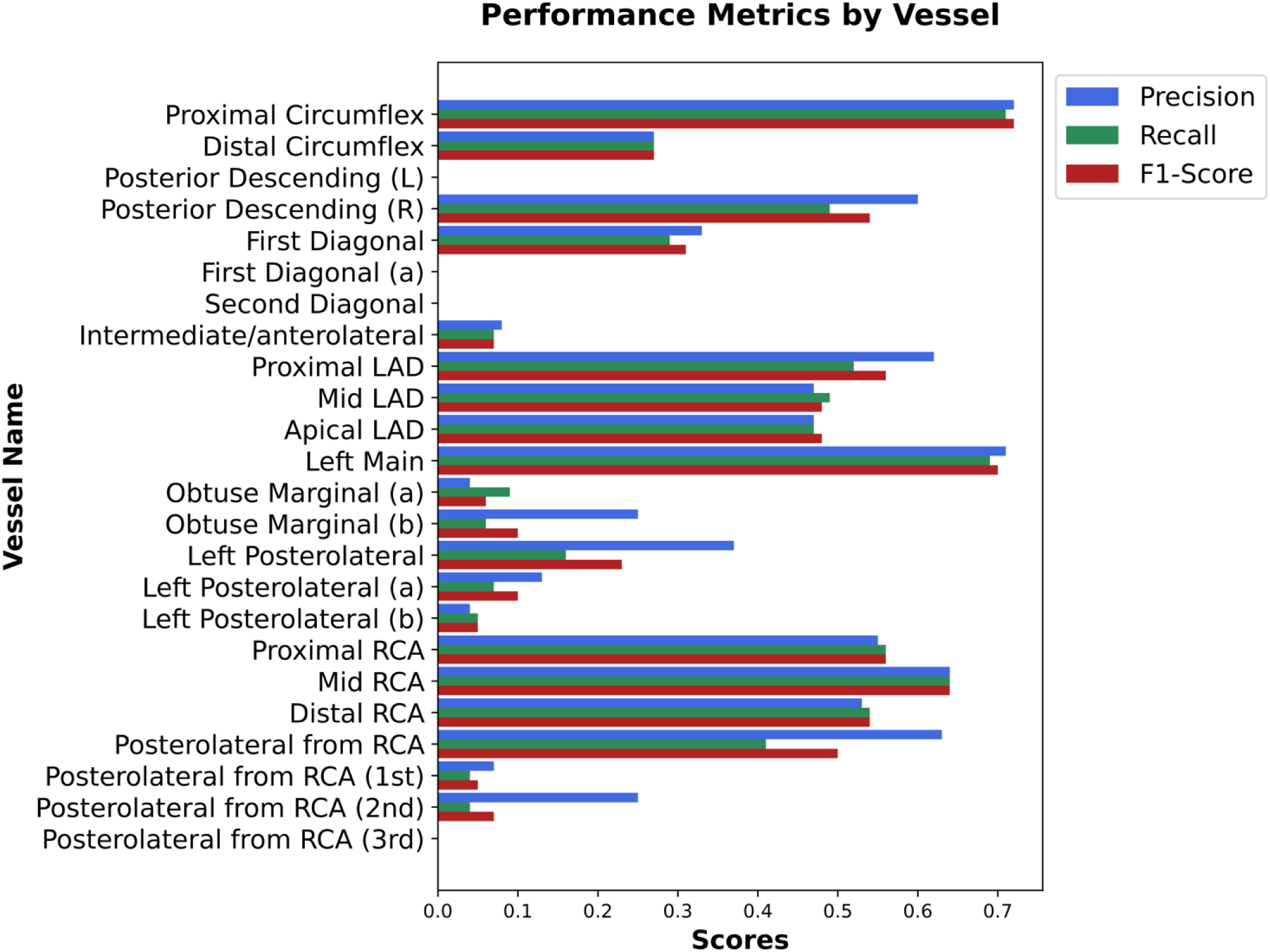
Bar chart showing performance of fine-tuned InternVL2-4B on anatomy labelling. Higher scores were seen in major vessel segments such as the left main, main branches of the circumflex, LAD and RCA, which were represented in higher proportion in the training set. Performance on the proximal segments were also better compared to the distal ones. Conversely, predictions on vessels that were less represented in training set tend to perform worse.

### 3.4 Report Generation

The report generation performance metrics of the fine-tuned InternVL2-4B is summarised as a heatmap in Fig 4. The VLM could generate text reports in a structured and standardised format. However, the correctness of the findings was less than satisfactory. In particular, it had high false positive (0.48) and negative rates (0.77), with low precision (0.19) and recall (0.23). It was observed that the VLM output no clinically significant stenosis on the left main coronary artery segment for all test cases, even though five presented with clinically significant ones. Additionally, the VLM also hallucinated in some test cases, describing the presence of collaterals or anatomical variations when there were not any. Some of these descriptors used were the same phrases that appeared in training data, suggesting a learning problem. These findings suggest that the VLM struggled to learn effectively from the training data, and a detailed explanation is offered in the next section.

**Fig 4.**
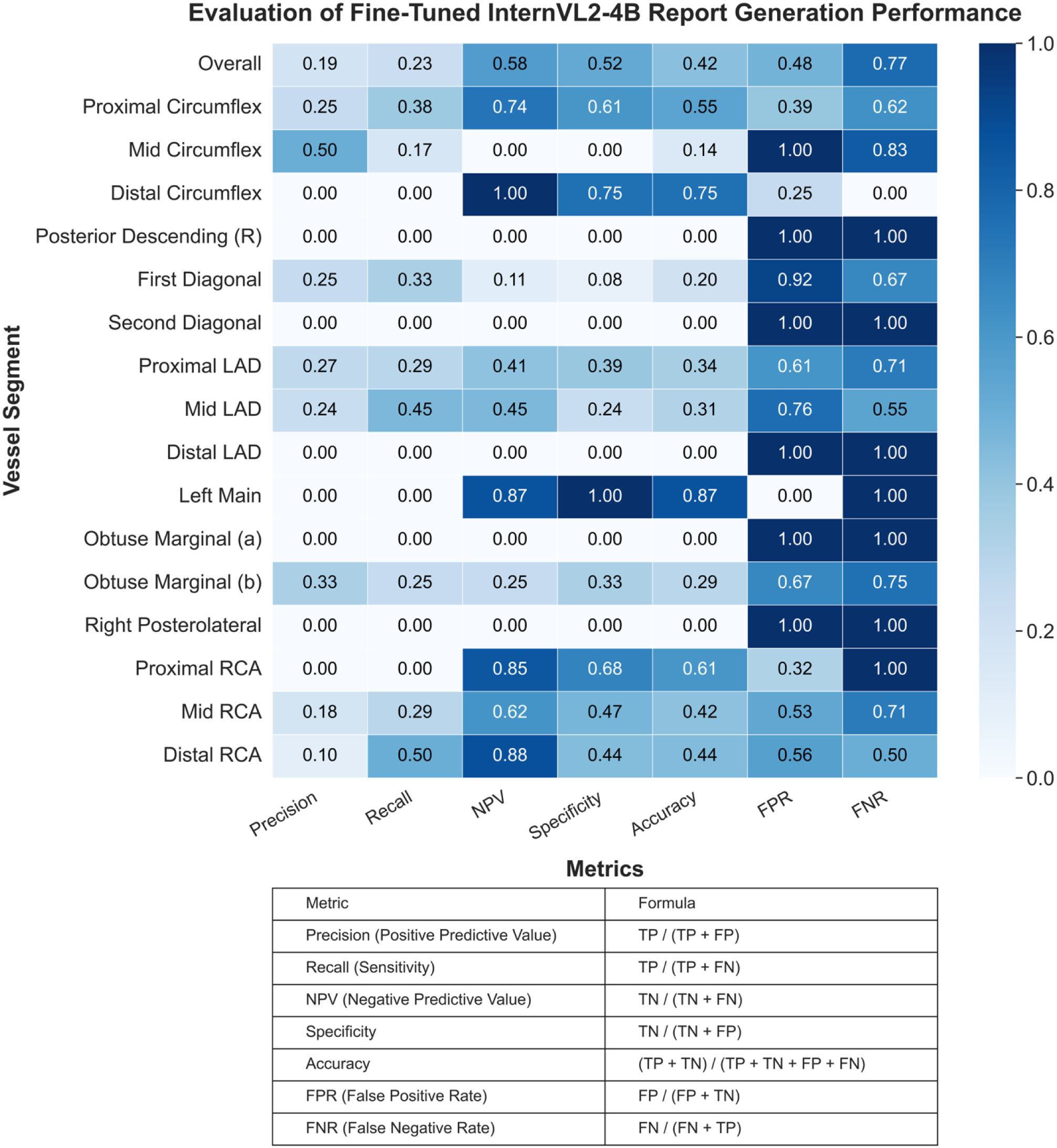
Heatmap showing the performance metrics of InternVL2-4B on report generation. The model was evaluated for its ability to detect stenosis and classify severity for each coronary vessel segment.

## 4 DISCUSSIONS

In summary, we trained an effective ViT keyframe selector and a VLM with strong performance in stenosis detection and anatomy labelling, but poor performance in report generation. Although the keyframe selector demonstrated robust performance, a major limitation in its application is the lack of clinical validation by an experienced cardiologist. Furthermore, the training data was prepared based on the assumption that all selected frames from the open-source datasets were indeed keyframes. However, this may not be true as a minority of the CADICA and ADSD datasets contain some frames in which only the proximal segment of the vessels were filled by contrast. These images were not filtered and were retained as key frames for training to maintain dataset completeness and consistency.

Additionally, while IoU is one of the most commonly used metrics in object detection for image processing, it has several limitations in evaluating stenosis detection. One major issue is the lack of standardisation in stenosis annotation. For example, a diffuse stenosis along a vessel can be annotated using either a single large bounding box, or multiple smaller bounding boxes. This difference could fundamentally influence how the VLM learns during training, and substantially affect the IoU calculation during evaluation. The annotation protocols are not explicitly standardised in the three datasets we used, leading to several inconsistencies. Furthermore, in CADICA and ADSD, bounding boxes tend to be smaller than those in the ARCADE dataset (Table 1). This suggests that these two datasets had more focal stenoses, while ARCADE curated more diffuse stenoses. It was also observed that some stenoses may not have been annotated in the ground truth. These lesions may be erroneously marked as FPs if detected by the VLM. As such, a qualitative assessment of the VLM is also crucial in objectively evaluating its performance. Fig 5 illustrates how metrics like IoU tend to underestimate the true performance of the VLM using examples from test cases. Lastly, there is also a disproportionate number of images with stenosis (19,522) compared to those without (2,130), resulting in a skewed training set. All test images contained diseased vessels, which means the ability of our VLMs to recognise non-diseased angiograms could not be accurately assessed. This imbalance in training dataset could also result in high false positive rates in a real-world scenario, and future work should aim to introduce more normal angiogram images into the training set.

**Fig 5.**
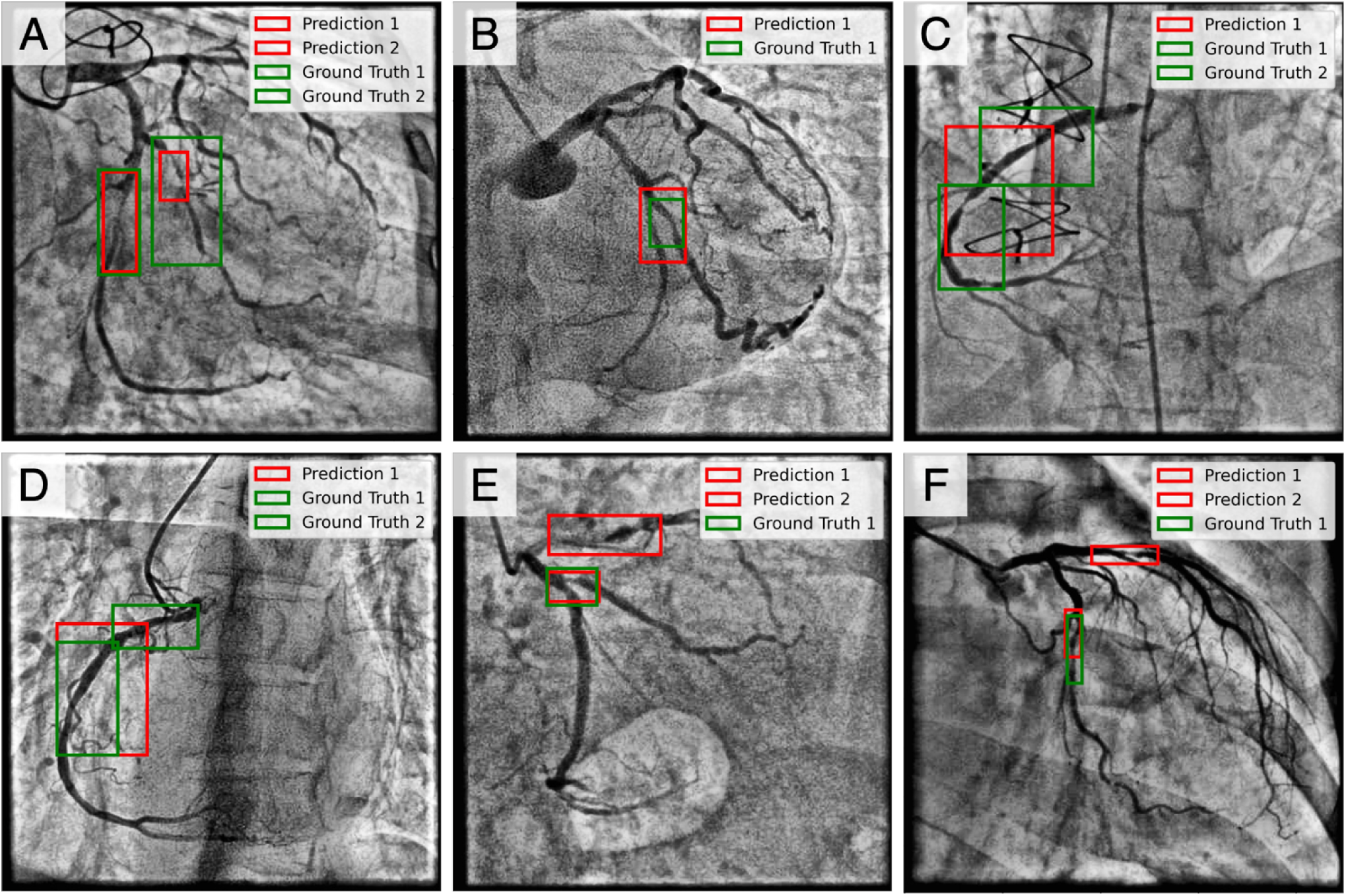
Selected examples of test cases on stenosis detection. (A) Prediction 1 (left) overlaps well with the corresponding ground truth with IoU of 0.77, is classified as TP. Prediction 2 (right) has low IoU of 0.15 with the ground truth. However, it is still classified as a TP since more than 60% of its area falls within the ground truth. (B) Prediction 1 has a larger area than the ground truth. Even though it correctly localises the stenosis, its IoU of 0.47 with ground truth is lower than the threshold. Hence, it is not matched to the ground truth, leading to a FP and a FN. (C) This angiogram shows a diffuse stenosis with regions of focal stenoses on the RCA. The ground truth annotates the stenoses using two bounding boxes. Prediction 1 has low IoU of 0.24 with both ground truths. Even though it localised stenosis correctly, this image has one FP and two FNs. (D) Prediction 1 matches ground truth 1 (left) with high IoU of 0.58. However, part of this prediction also accounts for stenoses annotated in ground truth 2 (right). However, the algorithm would consider ground truth 2 as unmatched. This results in 1 TP and 1 FN for this image. (E) Prediction 1 (below) is TP as it overlaps well with ground truth 1 with IoU of 0.75. However, the VLM also generated prediction 2 (above), which indicates a possible stenosis that was not annotated in the ground truth. Prediction 2 is marked as a FP in this case. (F) Prediction 1 (left) is a TP as >60% of its area overlaps with the ground truth. However, prediction 2 indicates a possible stenosis that was not annotated in the ground truth, leading to a FP.

In anatomy labelling, segmentation at the pixel level would be more precise as the area bounded by the bounding boxes tends to contain more background than the vessel segments due to differences in their shapes. However, we opted to experiment with a more resource-efficient approach by using the InternVL2-4B model instead of larger InternVL variants that support pixel-level segmentation but require substantially more GPU capacity. When fine-tuned, the InternVL2-4B VLM demonstrated strong performance in basic coronary artery anatomy labelling tasks, though the results are not directly comparable to those from existing studies. Future work should incorporate more training images focusing on these smaller vessel segments to improve model performance. If computational resources allow, fine-tuning using larger models such as InternVL2-26B can be experimented to study vessel labelling at the pixel level for better performance.

The poor performance in report generation could be attributed to several resource limitations in the methodology of this study. During the training for report generation task, each training sample consists of multiple angiographic images paired with a single consolidated text report without explicit annotations or breakdowns for each image. This could have resulted in high information density within the training example, as the model struggles to infer multiple diagnostic insights from limited supervised labels and no explicit reasoning guidance. This problem is exacerbated by limited training data, which only contained about 300 cases. Exploring the attention layers in future studies may provide valuable insights into how the images are being interpreted, thereby enhancing the model’s explainability. This analysis was not conducted in the present work, as it was considered beyond the scope of this preliminary study. Given that the fine-tuned InternVL2-4B performed well on single-image tasks like stenosis detection and anatomy labelling but did poorly in multi-image-based tasks such as full report generation, the complexity of multi-image analyses is a highly probable cause for the poor performance. A logical next step would be to pair diagnostic findings to each image rather than the full set of images collectively. However, this would require some specialised clinical knowledge and time to annotate images individually, which unfortunately was not available at the time of this study.

## 5 FUTURE DIRECTIONS

In this study, we attempted to generate angiogram reports by training and testing InternVL2-4B to read multiple selected angiogram frames and generate a full diagnostic report. However, there was no explicit guidance on how each image should be interpreted. In clinical practice, each coronary artery segment has a standard projection angle in which lesions are most optimally visualised. However, due to time constraints, it was not possible to have annotations for every individual image. Instead, all images belonging to the same patient were grouped and paired to the diagnostic report for training and testing. This has resulted in a weakly-supervised learning problem, worsened by small sample size. An immediate and logical next step would be to annotate these images individually by pairing only relevant portions of the report that accurately describe the image. This can significantly reduce the complexity for the VLM during learning as it improves the precision of supervision.

Additionally, since the AIMx dataset contains raw video files in the DICOM format, metadata information such as cranial-caudal and left/right-anterior oblique projection angle values can be supplemented to every image to guide the VLM in reasoning out which segment is being visualised. These improvements would maximise the advantages of VLMs for coronary angiogram interpretation tasks, bringing it closer to real-world application.

## 6 CONCLUSIONS

This study is the first to explore using VLMs for interpreting coronary angiograms and generating a comprehensive diagnostic text report by integrating multiple angiographic projections. Although the current capabilities of the VLM are limited in terms of full report generation, we have identified several probable and addressable factors that future studies should refine to enhance its performance. We have also demonstrated InternVL2-4B’s strong capability in stenosis detection and anatomy labelling after finetuning with LoRA, underscoring the significant potential of VLMs in automating coronary angiogram interpretation and clinical implementation. This study has laid a strong foundation as a proof of concept by demonstrating its feasibility, and has identified key limitations which could be resolved with continued refinement.

An accurate and reliable AI model that can interpret coronary angiograms has diverse applications. In the clinical setting, it could serve as a visual aid to cardiologists by annotating areas of stenosis and important vessel segments. It would also improve clinical workflow by reducing administrative time for report generation and computation of the SYNTAX score. Such a model could improve diagnostic accuracy, especially in resource-limited settings like developing countries or regions where general cardiologists instead of specialised interventionalists perform diagnostic angiography. In medical education, it could serve as an interactive learning tool, providing trainees with feedback on lesion identification and anatomy recognition using real angiogram images. Beyond the clinical space, an AI model capable of angiogram interpretation could also benefit stakeholders such as regulatory bodies and insurance companies. It can be utilised to audit medical decisions, ensuring adherence to medical guidelines by identifying potential malpractices such as unnecessary percutaneous coronary interventions for clinically insignificant stenosis. Given its diverse applications, further investment in developing such a model is well justified.

## 8 DATA AVAILABILITY

The AIMx dataset and model weights used in this study are not publicly available due to patient privacy concerns but may be made available from the corresponding author on reasonable request, according to the policies from the SingHealth Institutional Review Board. Other datasets used in this study are open-source and can be directly obtained from the references.

## 9 CODE AVAILABILITY

The evaluation scripts for stenosis detection and anatomy recognition are available at: https://github.com/jiang-qf/VLM-for-Coronary-Angiography.

## 11 AUTHORS’ CONTRIBUTIONS

Conceptualization and guidance: Y.H. Ke, L.G. Sinisterra, K.K. Yeo, Y. Jonathan, D.S.W. Ting. Coding and technical development: Q. Jiang, K. Elangovan, Z. Li.

Data analysis: Q. Jiang, L.G. Sinisterra, K. Elangovan, Z. Li. Manuscript preparation & proof-reading: All authors

